# Multimodal Fusion of Pathology Free-Text and Clinical Data Enhances Complication-Risk Discrimination After Implant-Based Breast Reconstruction

**DOI:** 10.64898/2026.01.20.26343695

**Authors:** Yimeng He, Hamzah Almadani, Sunny Huang, Judith Monzy, Debiao Li, Edward Ray, Xiuzhen Huang

**Affiliations:** Department of Computational Biomedicine, Cedars-Sinai Medical Center, Los Angeles, California 90048, USA; Biomedical Imaging Research Institute, Cedars-Sinai Medical Center, Los Angeles, California 90048, USA; Department of Surgery, Cedars-Sinai Medical Center, Los Angeles, California 90048, USA

## Abstract

Implant-based breast reconstruction is the most common surgical option following mastectomy for breast cancer. Despite its prevalence, up to one-third of patients develop complications within two years. Existing machine-learning models for predicting the complications rely solely on structured clinical data, overlooking prognostic information in narrative pathology reports. Recent advances in large language models (LLMs) enable extraction of numeric and semantic information from clinical text, offering opportunities to improve predictive performance and interpretability. We developed a fully on-premises, open-source model that extracts numeric morphometrics and contextual embeddings from free-text pathology reports, and fuses them with 63 structured variables via a CLIP-style dual encoder. In a single-center cohort of 963 patients (Jan 2007–Jan 2022), the multimodal model improved composite-complication risk discrimination (AUROC from 0.691 with logistic regression, increased to 0.740 with clinical features, and further improved to 0.764 with the addition of pathology report text features; p=0.027) and enhanced sensitivity and positive predictive value at clinical thresholds. The automated extraction module we developed for numeric morphometrics (e.g. mastectomy-specimen weight) from free-text pathology reports achieved an accuracy of 96.3%. SHAP analyses confirmed established risk factors—expander-to-implant interval, body-mass index, and total mastectomy weight—as dominant drivers. In subgroup analyses, model performance remained robust, with particularly strong discrimination observed among specific populations, such as shorter expander-to-implant interval (EII), the AUROC reached 0.796 and accuracy was 0.799.

These results show that on-premises, open-source LLMs reliably extract and fuse textual and structured clinical features to achieve clinically meaningful gains in predicting complications after implant-based breast reconstruction. While traditional models are constrained by the limited scope of structured variables, pathology text—when analyzed with modern language models—adds new, clinically relevant signals. Even modest statistical gains yield more accurate identification of high-risk patients, potentially informing surgical planning, patient counseling, and postoperative follow-up. These findings demonstrate that privacy-preserving language models, when integrated with contrastive multimodal alignment, can unlock prognostic information embedded in narrative pathology reports and enable interpretable, patient-level decision support. This interpretable, privacy-preserving multimodal framework offers a generalizable approach for enhancing risk prediction and clinical decision-making across surgical oncology.

**Significance Statement:** This study demonstrates that multimodal fusion of pathology free-text and structured clinical data, enabled by on-premises large language models, improves complication-risk discrimination after implant-based breast reconstruction. The approach uncovers prognostic signals hidden in narrative reports, offering an interpretable, privacy-preserving framework for precision risk prediction in surgical oncology.

## 1 Introduction

Breast cancer is the most commonly diagnosed cancer in women worldwide [1–3]. Mastectomy for breast cancer is often followed by breast reconstruction; large U.S. cohorts report that roughly 40–60 % of women undergo breast reconstruction within two years of surgery, with utilization rising steadily over the past two decades [4, 5].Implant-based techniques outnumbering autologous flaps by roughly three-to-one [2, 3], largely because they halve operative time, shorten hospital stay and eliminate donor-site morbidity, albeit at the cost of higher early device-related complications [6, 7] Large multi-center cohorts show that around 27–33 % of women experience at least one adverse event—most commonly infection, seroma, malposition or device loss—complication within two years of post-mastectomy reconstruction [8, 9].Complication rates vary further with timing (immediate vs delayed) and staging (direct-to-implant vs tissue-expander), complicating shared decision-making and underscoring the need for reliable, patient-specific risk stratification.

Despite steady interest, every published machine-learning model for implant-based breast-reconstruction complications still relies *solely* on hand-curated, structured variables. In the largest series to date, Chen *et al*. [10] reached an AUROC of 0.74 with a feed-forward neural network, while Hassan *et al*. [11] and Naoum *et al*. [12] reported values of around 0.73 using ensemble and random-forest approaches.

Transformer large-language models can now extract categorical findings from clinical narratives with *>* 95% accuracy [13, 14], yet *numeric* morphometrics such as mastectomy-specimen weight remain untouched, and no study has fused any text-derived features with structured clinical data. Thus, the information-rich free-text pathology report remains an untapped modality, leaving a clear gap in current risk-prediction frameworks.

The recent proliferation of high-performing, *open-source* LLMs—for example Gemma 3 and Llama 3—now permits hospital-firewalled, cost-free natural-language processing at near–state-of-the-art accuracy [15, 16]. Concurrently, contrastive pretraining frameworks such as CLIP have demonstrated strong ability to align heterogeneous modalities and to drive medical-AI benchmarks when adapted for domain-specific text–image or text–signal tasks [17, 18]. These advances raise an untested question: can automatic extraction of numeric morphometrics and contextual embeddings from free-text pathology reports, when fused with structured clinical variables via a CLIP-style dual encoder, improve complication-prediction beyond tabular-only baselines? We address this question by combining on-premises LLM extraction with a multimodal, contrastively trained predictor and assessing its performance against the best existing structured-data models.

Here we show that this fully on-premises, multimodal pipeline achieves 96.3 % extraction accuracy (Wilson 95 % CI 89.7–98.7) and lifts composite-complication prediction after implant-based reconstruction from an AUROC of 0.691 to 0.764 while retaining SHAP-interpretable feature attributions.

## 2 Results

### 2.1 Cohort assembly

We assembled a single-center cohort of consecutive patients undergoing implant-based breast reconstruction between Jan 2007 and Jan 2022. We linked de-identified pathology reports to the clinical registry and restricted to patients present in the implant registry. Inclusion required an available pathology report and target outcome label. Exclusions were prespecified: implausible expander-to-implant interval (`TEtoImplant`) outside 1 year (See Fig 1 for details).

**Fig. 1:**
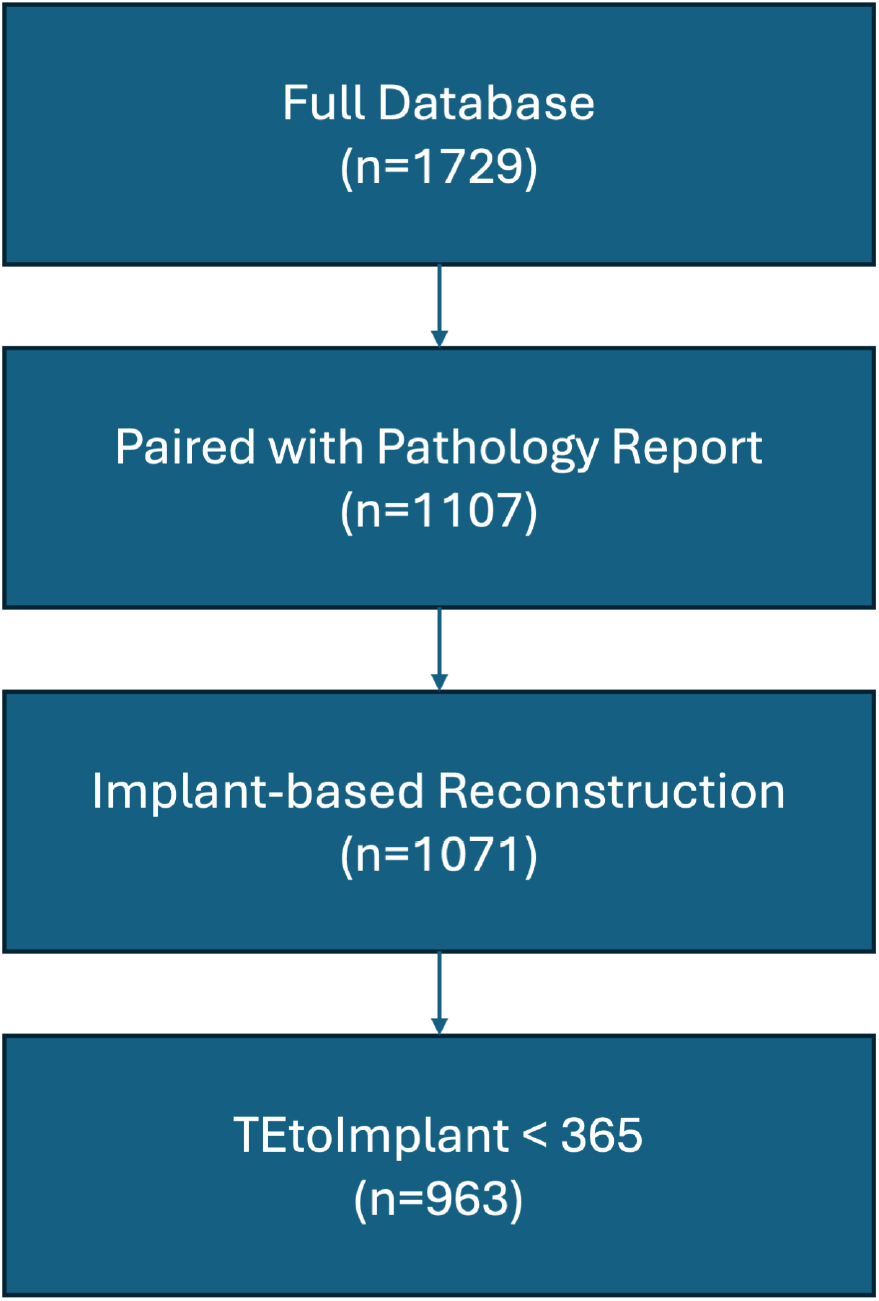
Cohort derivation. Starting from the full registry (*n* = 1,729), cases were linked to an available pathology report (*n* = 1,107) and restricted to implant-based reconstruction procedures (*n* = 1,071). The analytic cohort comprised *n* = 963 patients after excluding records with an expander-to-implant interval `TEtoImplant` ≥ 365 days. `TEtoImplant` denotes days from tissue expander placement to implant exchange.

**Fig. 2:**
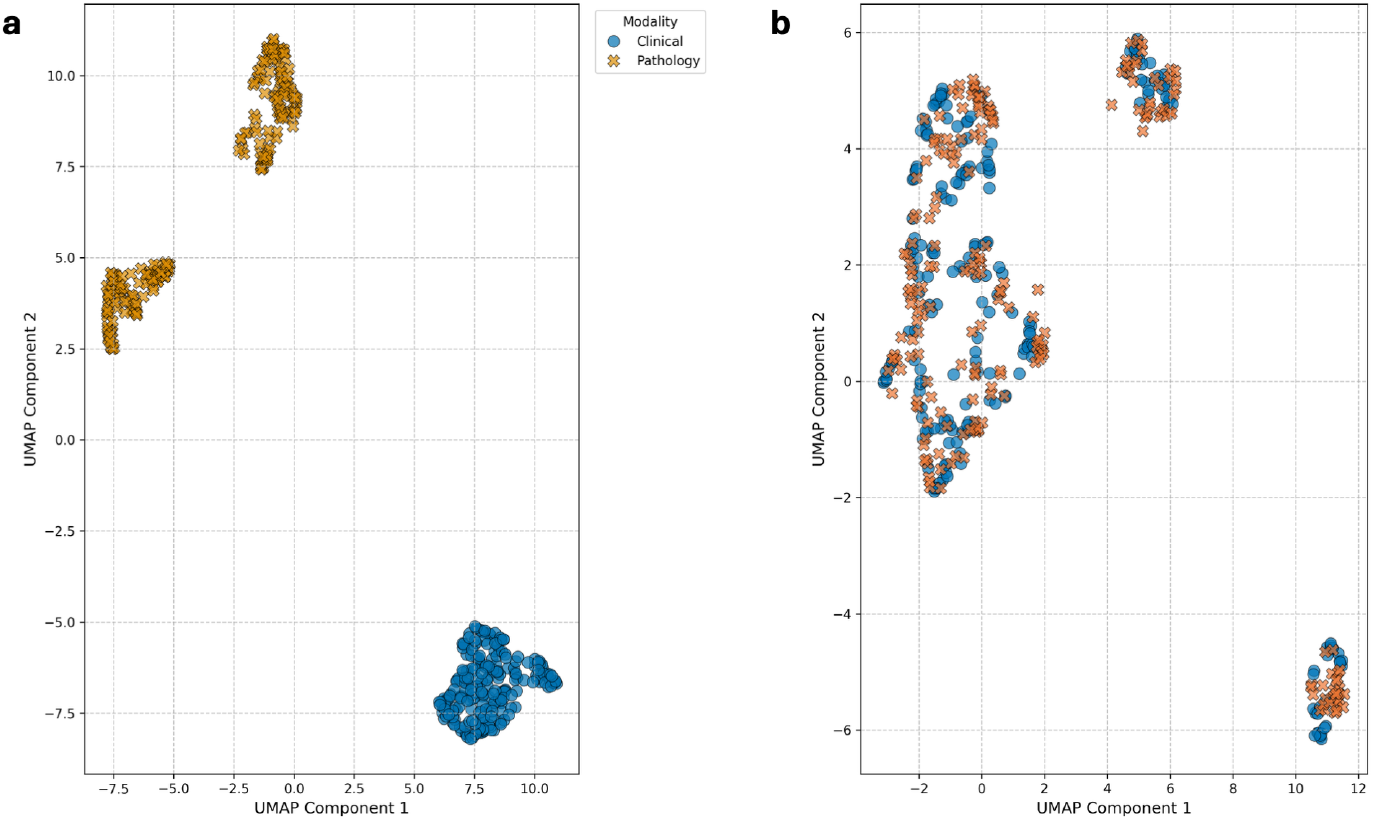
UMAP visualization of Clinical and Pathology data embeddings before and after CLIP model training. (a) Before CLIP training, the UMAP projection shows Clinical (blue circles) and Pathology (orange x-marks) data modalities forming distinct and well-separated clusters, indicating no initial alignment in this embedding space. (b) After CLIP training, the UMAP projection demonstrates significant intermingling of Clinical (blue x-marks) and Pathology (orange circles) data. This mixture indicates that the CLIP model has successfully learned a shared repre-sentation space where features from the two modalities are aligned.

963 women underwent implant-based breast reconstruction at Cedars–Sinai Medical Center (CSMC) and were included in this analysis. The mean age was 56.3 ± 11.6 years and the mean body-mass index (BMI) was 24.9 ± 4.8 kg/m^2^. Diabetes mellitus and hypertension were present in 6.6% and 16.9% of patients, respectively, and the overall incidence of implant-based complication was 30.5% (Table S1).

### 2.2 LLM extraction is accurate and failure modes are predictable

The performance of the open-source Gemma 3 LLM in extracting breast specimen weight from pathology reports was evaluated by comparing its output against measurements manually extracted by a medical doctor. From an initial cohort of 60 patients, a total of 37 left breast reports and 44 right breast reports contained sufficient information for a valid comparison of extracted weights. (Table 1)

**Table 1:** Accuracy and 95% Wilson Score Confidence Intervals for Breast Weight Extraction.

| Dataset | Correct/Total | Accuracy (%) | 95% Wilson CI (%) |
| --- | --- | --- | --- |
| Left | 37/37 | 100.00 | [90.59, 100.00] |
| Right | 41/44 | 93.18 | [81.77, 97.65] |
| Combined | 78/81 | 96.30 | [89.67, 98.73] |

For the left breast, the model achieved perfect accuracy (100%, 37 out of 37 cases correctly extracted; 95% Wilson CI: 90.59% - 100.00%) when compared to the physician’s manual extraction.

For the right breast, the model demonstrated an accuracy of 93.2% (41 out of 44 cases correctly extracted; 95% Wilson CI: 81.77% - 97.65%). The 3 instances of incorrect extraction on the right side all shared a common pattern: the pathology report did not explicitly mention the mastectomy specimen weight, and the model erroneously extracted the weight of the breast implant as the specimen weight. In all other 41 cases for the right breast where specimen weight was available, the model extracted it correctly.

### 2.3 Clinical and pathology text align in a shared space

#### 2.3.1 Cross-modal alignment

Without contrastive alignment, clinical (blue) and pathology (orange) vectors occupy disjoint regions (Fig.2a). After training with CLIP loss and an auxiliary laterality classifier, the two modalities intermingle in the same UMAP manifold (Fig.2b), indicating patient-level correspondence.

We evaluated alignment on the 215-pair validation set. From clinical → pathology, Recall@K was *R*@1 = 0.074, *R*@5 = 0.288, *R*@10 = 0.409, *R*@20 = 0.581, and *R*@50 = 0.781, corresponding to 16.0×, 12.4×, 8.8×, 6.25×, and 3.36× enrichment over a random baseline (*K/N*, *N* =215). The reverse direction (pathology → clinical) was similar (*R*@1 = 0.088 [19.0×], *R*@5 = 0.279 [12.0×], *R*@10 = 0.414 [8.9×], *R*@20 = 0.554 [5.95 ×], *R*@50 = 0.772 [3.32×]). Although absolute top-*K* recalls are moderate on this small validation set, the aligned model retrieves the correct cross-modal counterpart substantially more often than chance.Table S2).

#### 2.3.2 Semantic retention

We froze the encoder weights of our `clip` model and trained lightweight logistic-regression probes on the frozen `[CLS]` embeddings (validation set, *n*=215). For predicting laterality (*unilateral* vs. *bilateral*), we used the pathology `[CLS]` and obtained AUROC 0.976 ± 0.027. For the remaining attributes—No Prior Radiation Therapy, prior smoker status, and hypertension status—we used the clinical `[CLS]`. Performance was high across attributes (Table 2): No Prior Radiation Therapy achieved 1.000 (SD undefined; perfect separation on the validation set), prior smoker status reached 0.977 ± 0.014, and hypertension status remained detectable at 0.853 ± 0.082. These results indicate that the `[CLS]` embeddings not only support cross-modal alignment but also retain granular clinical semantics that are recoverable with simple linear probes.

**Table 2:** Discriminative performance of auxiliary classifiers on frozen [CLS] embeddings from the CLIP model (validation set, *n* = 215). Laterality was predicted using the pathology [CLS] embedding, while other clinical attributes were predicted using the clinical [CLS] embedding.

| Attribute | AUROC (SD) |
| --- | --- |
| Laterality (unilateral vs. bilateral)<sup>a</sup> | 0.976 (27) |
| No prior radiation therapy<sup>b</sup> | 1.000 (—) |
| Prior smoker status | 0.977 (14) |
| Hypertension status | 0.853 (82) |

<sup>a</sup> Laterality predicted using the *pathology* [CLS] embedding; other attributes use the *clinical* [CLS] embedding.
<sup>b</sup> SD undefined (perfect separation); shown as an em dash. Values in parentheses denote SD in absolute units; e.g., (27) = 0.027.

### 2.4 Multimodal fusion yields consistent gains in discrimination

As an anchor, multivariable logistic regression with identical preprocessing achieved AUROC 0.691 ± 0.054 (L2) (Table S2). Benchmarking on the same 62 structured clinical variables, the Clinical LightGBM improved discrimination to 0.740 ± 0.050 across ten stratified folds (Δ = +0.049 vs. L2). For meaningful head-to-head comparison, we designate this Clinical LightGBM as the baseline for the remainder of the paper—it was the best-performing clinical-only model in cross-validated benchmarking—and all reported Δ values are computed relative to this baseline (Table 3). Augmenting the baseline with morphometrics extracted from the pathology report—most notably total mastectomy-specimen weight—yielded only a marginal change (AUROC 0.744±0.039; Δ = +0.004 vs. baseline; paired Wilcoxon signed-rank *p* = 0.49).

**Table 3:** Cross-validated discrimination of complication-prediction models (*n* = 10 folds).

| Model / Feature set | AUROC (SD) | ΔAUROC<sup>a</sup> | p-value<sup>b</sup> |
| --- | --- | --- | --- |
| Baseline (Clinical LightGBM) | 0.740(50) | +0.049 | — |
| Baseline + LLM extracted breast weight (M2) | 0.744(39) | +0.004 | 0.492 |
| **Ensemble models (class weights 1:1)** |  |  |  |
| M2 + [CLS]<sub>clinical</sub> | 0.758(58) | +0.018 | 0.064 |
| M2 + [CLS]<sub>pathological</sub> | 0.760(60) | +0.020 | 0.105 |
| M2 + [CLS]<sub>both</sub> | **0.764(58)** | **+0.024** | **0.027\*** |

<sup>a</sup> $\Delta$ AUROC relative to Baseline.<sup>b</sup> Paired Wilcoxon signed-rank test.
Values in parentheses denote the standard deviation (e.g., (50) = 0.050).

We next constructed soft-voting ensembles by combining the M2 (clinical features + morphometrics feature) logits with 768-dimensional `[CLS]` embeddings from the dual encoder. Adding the *clinical* `[CLS]` improved AUROC to 0.758 ± 0.058 (Δ = +0.018; *p* = 0.064), though this did not reach significance. Using the *pathological* `[CLS]` yielded AUROC 0.760 ± 0.060 (Δ = +0.020; *p* = 0.105). The strongest performance was consistently obtained when both embeddings were included, achieving AUROC **0.764** ± **0.058** (Δ = +0.024; *p* = 0.027).

Threshold-specific operating characteristics mirrored this pattern (Table 4). At 80 % specificity, sensitivity improved from 0.573 (baseline) to 0.609 with the best ensemble. With sensitivity fixed at 80 %, specificity rose from 0.431 to 0.482, and positive predictive value at a 0.50 threshold improved from 0.668 to 0.828.

**Table 4:** Threshold-specific performance of complication-prediction models (*n* = 10 folds).

| Model / Feature set | Sens@80% Spec | Spec@80% Sens | PPV@0.5 |
| --- | --- | --- | --- |
| Baseline (Clinical LightGBM) | 0.573 | 0.431 | 0.668 |
| Baseline + LLM extracted breast weight (M2) | 0.585 | 0.427 | 0.709 |
| **Ensemble models** |  |  |  |
| M2 + [CLS]<sub>clinical</sub> | 0.597 | **0.482** | 0.815 |
| M2 + [CLS]<sub>pathological</sub> | **0.617** | 0.431 | **0.828** |
| M2 + [CLS]<sub>both</sub> | 0.609 | 0.452 | **0.828** |

For completeness we also evaluated different models and a “zero-shot” baseline that replaces the trained pathology embedding with the raw 5376-dimensional vector output by Gemma 3. None of these alternatives surpassed the both `[CLS]` ensemble (Supplementary Table S2); all imputation variants performed at or below the tabular baseline, and the zero-shot Gemma model reached an AUROC of 0.750.

### 2.5 Shorter expander–to–implant interval and radiation during staged reconstruction yield the highest predictive performance

Using out-of-fold predictions from the best ensemble model, discrimination was similar by age: Age *<* 65 years (AUROC = 0.756; 95% CI 0.709–0.800) vs Age ≥ 65 years (0.770; 0.693–0.836; Fig. 3a). By contrast, performance varied with the expander–to–implant interval: *<* 180 days showed higher discrimination (0.796; 0.747–0.837) than ≥ 180 days (0.620; 0.532–0.705; Fig. 3b). For radiation timing, AUROC was highest during staged reconstruction (0.798; 0.714–0.877), followed by before reconstruction (0.767; 0.626–0.886) and no radiation (0.731; 0.682–0.781), with after reconstruction lower (0.587; 0.397–0.761; Fig. 3c). Across panels, confidenceinterval overlap suggests minimal age effect, whereas interval and timing exhibit clear separations; wider CIs in smaller strata (e.g., post-reconstruction) warrant cautious interpretation.

**Fig. 3:**
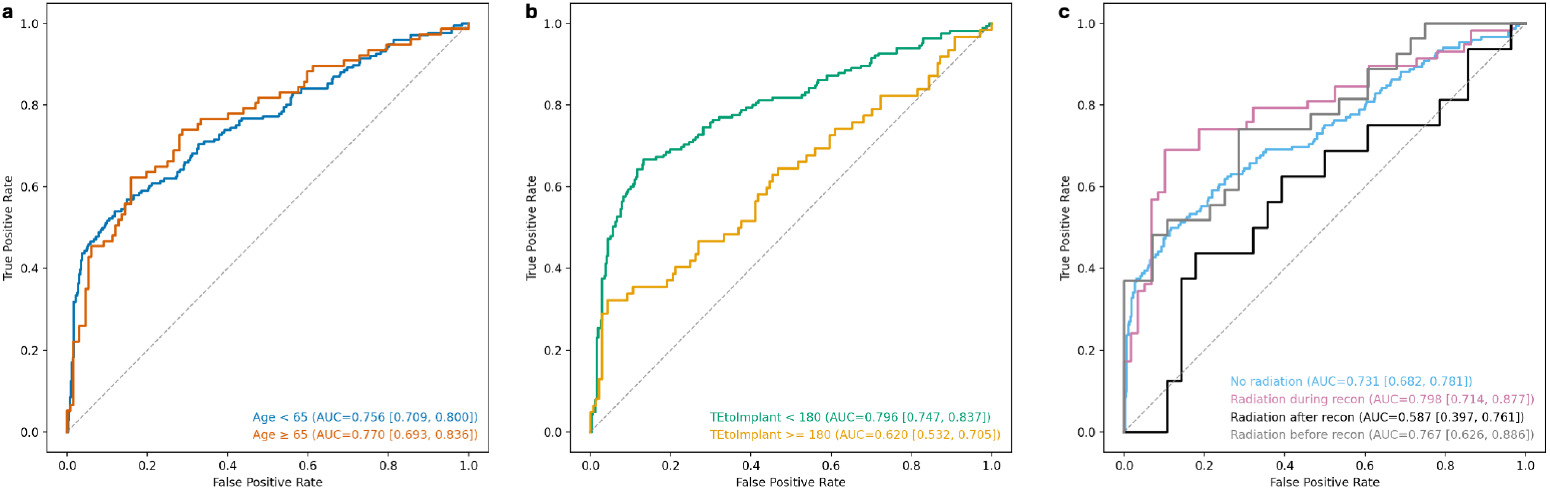
Subgroup ROC curves using out-of-fold predictions from the best model. **(a)** Age strata show similar discrimination: Age *<* 65 (AUC= 0.756; 95% CI 0.709–0.800) vs. Age ≥ 65 (AUC= 0.770; 0.693–0.836). **(b)** Expander–to–implant interval (EII; `TEtoImplant`) shows higher discrimination when *<* 180 days (AUC= 0.796; 0.747–0.837) versus ≥ 180 days (AUC= 0.620; 0.532–0.705). **(c)** Radiation timing: during staged reconstruction yields the highest performance (AUC= 0.798; 0.714–0.877), followed by before reconstruction (AUC= 0.767; 0.626–0.886) and no radiation (AUC= 0.731; 0.682–0.781); after reconstruction is lower (AUC= 0.587; 0.397–0.761). Dashed line denotes a no-skill classifier. 95% CIs were computed via a stratified patient-level bootstrap that resampled positives and negatives separately to preserve class balance.

Supplementary Table S4 summarizes other factors: AUROC was about 0.77–0.78 for hypertension, thyroid disorder, and hyperlipidemia, while the current-smoker subgroup showed a higher AUROC (0.847) but with limited sample size (*N* =31).

### 2.6 Attribution concentrates on timing, BMI, and specimen weight

Using SHAP on the LightGBM branch of the best ensemble, the top-10 features indicate that `TEtoImplant` contributes the largest share of predictive signal, followed by `BMI` and `total_weight` (Fig. 4). Smaller, but non-negligible, contributions are observed for `RadiationTiming_mid, Weight, Age, Mastectomy_doy, Height, RadiationTiming None`, and `HAS _HYPERTENSION`. The dependence plots (Fig. 5) show clear non-linear effects. For `TEtoImplant` (colored by `BMI`), very short intervals (∼ 0– 30 days) are associated with strongly positive SHAP values, a trough appears around ∼ 60–130 days, a secondary rise is visible near ∼ 150–210 days, and effects become increasingly negative beyond ∼ 250 days. For `total_weight` (colored by BMI), the pattern is approximately inverted-U: mid-range weights (∼ 500–1200 g) yield positive SHAP values, whereas very high weights (≳ 1800–2000 g) contribute negatively. Across both panels, the color gradient suggests that BMI *modulates* these effects only mildly; the primary non-linear relationships of `TEtoImplant` and `total_weight` dominate the contribution to the model output.

**Fig. 4:**
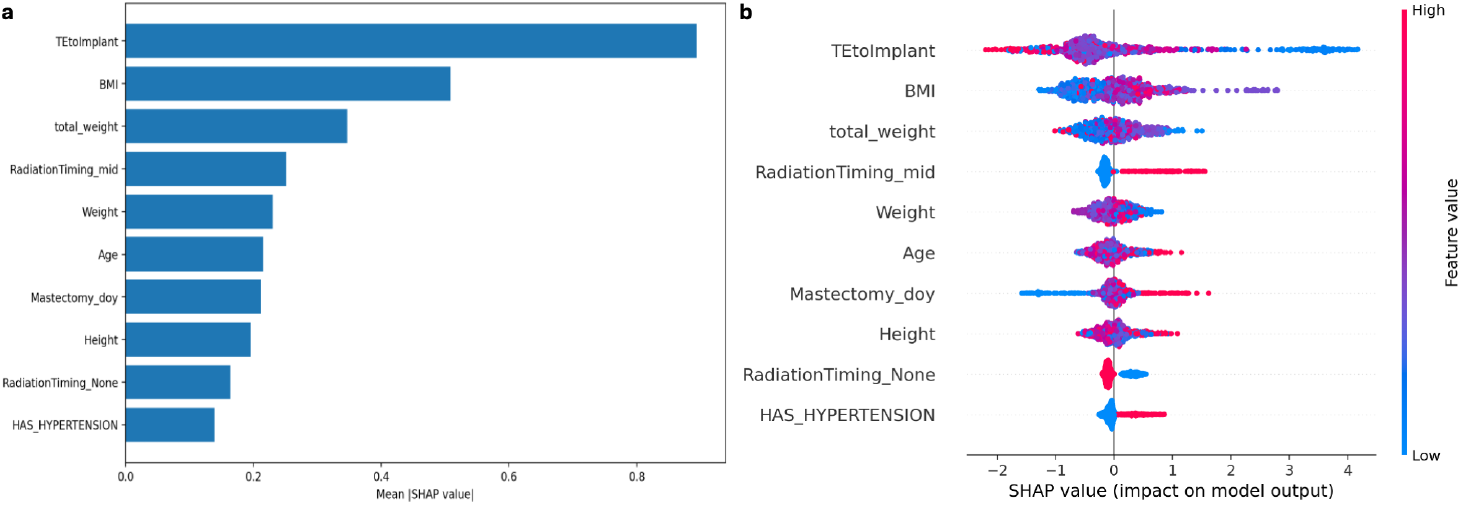
Top-10 feature contributions for the LightGBM branch of the model with both CLSs. **(a)** Mean absolute SHAP values highlight `TEtoImplant` as the largest contributor, followed by BMI and `total _weight`; smaller but non-negligible effects are seen for `RadiationTiming_mid, Weight, Age, Mastectomy_doy` (day-of-year, a seasonality proxy), `Height, RadiationTiming_None`, and `HAS_HYPERTENSION`. **(b)** Beeswarm plot for the same top-10 features shows directionality and dispersion of effects; warmer colors indicate higher feature values. Positive SHAP values indicate increased model output toward the positive class.

**Fig. 5:**
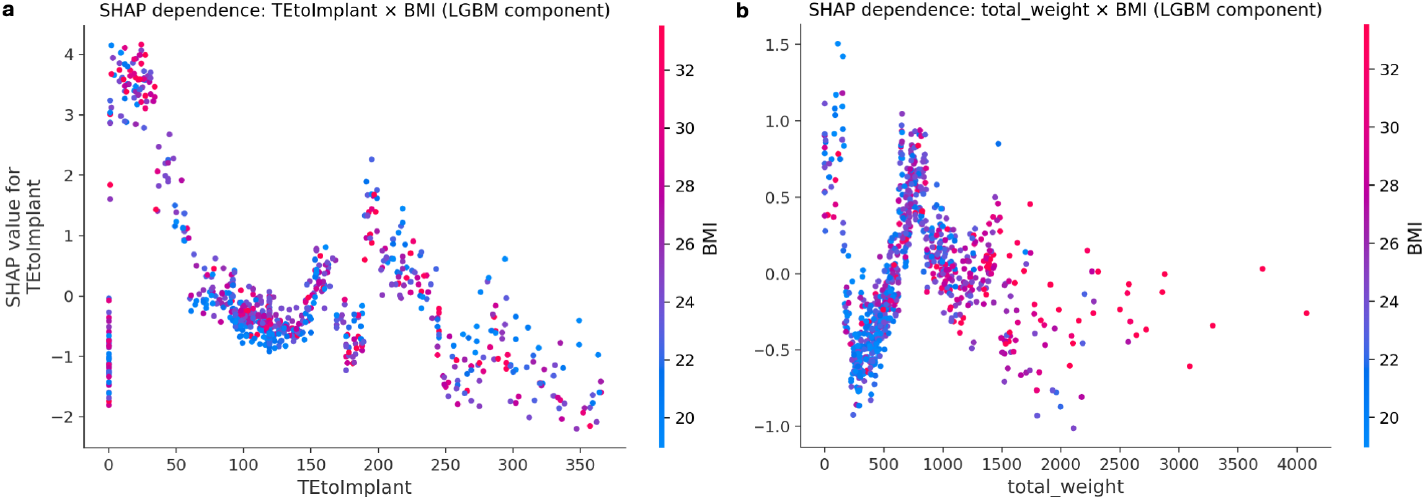
SHAP dependence plots from the LightGBM branch of the ensemble model. **(a)** `TEtoImplant` (days) vs. its SHAP value, colored by BMI (kg/m^2^). Short intervals (∼0–30 days) are associated with large positive SHAP values (greater probability of the positive class), a trough occurs around 60–130 days, a secondary rise appears near 150–210 days, and contributions become increasingly negative beyond ∼250 days. **(b)** `total_weight` (grams; sum of mastectomy specimen weights from the operated breasts) vs. its SHAP value, colored by `BMI`. An approximate inverted-U is evident: mid-range weights (∼500–1200 g) yield positive contributions, whereas veryhigh weights (≳1800–2000 g) are negative. In both panels, BMI modulates effects only mildly (color gradient) relative to the dominant non-linear main effects of `TEtoImplant` and `total_weight`. In all plots, a positive SHAP value indicates a shift toward the positive class.

## 3 Discussion

This study demonstrates that narrative pathology can be converted into reliable, on-premises signals using foundation models. When fused with structured variables, these signals improve risk prediction after implant-based reconstruction. Using an opensource LLM to extract numeric morphometrics and a dual-encoder to align clinical and pathology text, we achieved high extraction accuracy and observed a modest but consistent improvement over a Clinical LightGBM baseline via soft-voting fusion of text embeddings. Subgroup analyses showed age-stable discrimination, with higher performance for shorter expander–to–implant intervals and during-reconstruction radiation. SHAP attributions were clinically coherent—expander–to–implant interval, BMI, and radiation timing ranked among the top contributors—supporting plausibility of the learned signals.

We prospectively evaluated the accuracy of extracting mastectomy-specimen weight from pathology reports with an on-premises, open-source LLM. Left-sided reports were perfectly extracted (37/37; 100.0%; 95% Wilson CI 90.6–100.0), right-sided reports reached 93.2% (41/44; 95% CI 81.8–97.6), and the combined accuracy was 96.3% (78/81; 95% CI 89.7–98.7). Achieving this level of extraction with a freely available model has practical and strategic implications. First, all inference was performed on-premises, ensuring that protected health information never left institutional control and obviating external data-sharing agreements. Second, the absence of per-call fees makes the pipeline affordable for resource-constrained centers, enabling equitable adoption. Third—and most importantly—because the model weights are fully accessible, the model can be fine-tuned locally to accommodate institution-specific report styles, rare tumor subtypes, or new clinical fields, which is difficult or impossible with closed commercial APIs. Prior LLM studies have focused largely on single-label classification (e.g., abdominal pain, stage, histologic diagnosis)[13, 14]; to our knowledge, no prior work has addressed the compound task of determining laterality and extracting the corresponding numeric mastectomy-specimen weight. Our 96.3% accuracy (Wilson 95% CI 89.7–98.7%) therefore provides the first evidence that an open-source LLM can solve this task reliably.

A Clinical LightGBM baseline on structured variables achieved AUROC 0.740 ± 0.050; adding the single numeric morphometric (M2) yielded 0.744 ± 0.039 (Δ=+0.004). Soft-voting ensembles that fuse the baseline with text embeddings improved discrimination: adding pathology `[CLS]` reached 0.760 ± 0.060 (Δ=+0.020), adding clinical-summary `[CLS]` reached 0.758 ± 0.058 (Δ=+0.018), and combining both `[CLS]` achieved the best overall AUROC 0.764 ± 0.058 (Δ=+0.024). At fixed operating points, sensitivity at 80% specificity rose from 0.573 (baseline) to 0.617 and PPV at a 0.50 threshold from 0.668 to 0.828. We hypothesize that these gains arise because distributed representations encode semantics that tabular 0–1 variables cannot: the pathology `[CLS]` appears to capture fine-grained lexical cues (e.g., histologic qualifiers, margin descriptors, negations, numeric magnitudes with units, cross-sentence references), while the clinical-summary `[CLS]` contributes temporal relations and linguistic qualifiers (e.g., “no prior radiation,” therapy chronology, hedging/uncertainty) and offers redundancy that can denoise inconsistent structured entries. Their complementary nature—reflected in their additive effect—suggests that embeddings provide clinically meaningful, partially non-overlapping signal beyond simple indicator variables, yielding small but consistent multimodal improvements.

The superiority of shorter expander–to–implant intervals and during-reconstruction radiation is consistent with reduced care-path variability and richer, temporally aligned documentation, whereas prolonged intervals and post-reconstruction radiation likely reflect heterogeneous pathways (adjuvant therapy, interim complications) and unmeasured confounding that erode discrimination. Age showed minimal effect, suggesting physiologic risk is captured by correlated covariates (e.g., BMI, hypertension) and clinical selection. These explanations are plausible but exploratory and warrant confirmation with external, time-anchored analyses.

SHAP ranked the EII as the dominant contributor, followed by BMI and total weight; radiation-timing and operative-timing proxies had smaller but nontrivial effects. Partial-dependence and SHAP interaction patterns suggested a threshold-like rise in risk beyond ∼ 6–12 months of EII, with EII × BMI interactions indicating that delay amplifies risk more in higher-BMI patients, while early exchange attenuates BMI/weight effects. These findings align with clinical intuition and highlight a modifiable lever (earlier exchange) and an interpretable interaction (EII × BMI) for counseling and triage.

There are limitations. This single-center study may not capture the diversity of report templates, abbreviations, and units used elsewhere; such domain shifts could degrade extraction accuracy and downstream performance. Our composite endpoint groups heterogeneous complications, suggesting that per-complication models may yield clearer mechanistic signals. Our CIs are derived from a stratified patient-level bootstrap of pooled out-of-fold predictions and thus quantify sampling uncertainty of the evaluation set but not variability from model retraining. Longer expander–to–implant intervals likely reflect heterogeneous care pathways (e.g., adjuvant therapy, delayed expansion, interim complications), introducing unmeasured confounding and potential covariate shift that can degrade discrimination. Priorities include multi-center external validation with domain-adaptive fine-tuning, addition of imaging to achieve a fuller multimodal representation, temporal modeling for dynamic risk updates, and pragmatic engineering to reduce compute requirements and support broad adoption.

## 4 Methods

### 4.1 Data Collection

Data were collected from all patients undergoing breast reconstruction after mastectomy for cancer or cancer prevention over a 15 year period, from Jan 2007 to Jan 2022, at Cedars-Sinai Medical Center, with at least 6 months of follow-up from the date of mastectomy.

### 4.2 LLM-based information extraction

We extracted structured morphometric fields from free-text pathology reports with the Gemma 3(27b) large language model in a zero-shot setting. Each report (de-identified plain text) was provided verbatim to the model together with a fixed instruction prompt (below). Inference ran data-parallel on 4× NVIDIA L40S GPUs. Decoding was deterministic (greedy/near-zero temperature) to ensure reproducible outputs.

#### Prompt

From the pathology report below, extract:

- `left breast mastectomy specimen weight (in grams)`,
- `right breast mastectomy specimen weight (in grams)`,
- `medial-lateral, superior-inferior, anterior-posterior dimensions (in cm) for each side,`
- `distance to the deep margin (cm) for each side,`
- `number of lymph nodes removed (count) for each side.`

~~~
If any field is not mentioned, return ‘nan’ for that field.
Return only a JSON object with exactly these keys:
{
   “left_breast_weight”: <number or nan>,
   “right_breast_weight”: <number or nan>,
   “left_breast_dims”: [ML, SI, AP] or [nan, nan, nan],
   “right_breast_dims”: [ML, SI, AP] or [nan, nan, nan],
   “left_deep_margin”: <number or nan>,
   “right_deep_margin”: <number or nan>,
   “left_num_lymph_nodes”: <integer or nan>,
   “right_num_lymph_nodes”: <integer or nan>
}
~~~

The returned JSON was parsed with strict schema validation. Numeric fields were coerced to *float* (weights, dimensions, margins) or *int* (lymph node counts). Units were normalized to grams and centimeters (e.g., “kg”→g, “mm”→cm) when explicitly present in the text; otherwise values were kept as given. Arrays `[ML, SI, AP]` were required to have length 3; malformed arrays or non-finite values were set to nan. We computed `total_weight` as the sum of left and right mastectomy specimen weights when available. If the model returned invalid JSON, we applied a lightweight repair (e.g., fix trailing commas) and re-parsed; unrecoverable cases were recorded as `nan` for the affected fields.

We only use `total_weight` for the downstream task, because we can not derive a reliable volume only with the dims, and there are too many nan in other extracted fields.

### 4.3 Pathology–clinical embedding alignment

Each patient’s structured record was first mapped with hand-written templates to yield a short clinical narrative (e.g. “Body-mass index is 28; tumor stage II; diabetes: no”), which was paired with the original pathology report. The two texts were encoded by independent `PubMedBERT-base` transformers[19]. When a source document exceeded the 512-token limit, it was split into overlapping windows (length 512, stride 128); each window produced its own `[CLS]` embedding, and these chunk-level representations were combined by a learnable self-attention pooling layer that assigned weights to the windows and returned a single, context-aware vector. The pooled embedding was then passed through a modality-specific linear projection to a 768-dimensional latent space, yielding one fixed-length representation for the pathology text and one for the templated clinical narrative. Training minimized a CLIP-style InfoNCE contrastive loss[20] (*τ* = 0.07) that maximized cosine similarity for matched pathology–clinical pairs within each mini-batch, alongside two auxiliary objectives: (i) a soft-max classifier on the pathology [CLS] to predict specimen laterality (left, right, bilateral) and (ii) a multi-label sigmoid classifier on the clinical [CLS] to reconstruct five key attributes (BMI category, smoking status, hypertension, menopausal status, ER/PR receptor status). The composite loss was *L* = *L*_InfoNCE_ + *λ*_lat_*L*_lat_ + *λ*_clin_*L*_clin_ with *λ*_lat_ = *λ*_clin_ = 0.1. Models were trained for 100 epochs with AdamW; training ran on 1 NVIDIA L40S GPUs.

### 4.4 Machine-learning model training and evaluation

We compared LightGBM, logistic regression, random forest, SVC, and MLP using nested stratified 10-fold cross-validation: an outer loop to estimate generalization and an inner loop to tune hyper-parameters using mean AUROC. All preprocessing (imputation, one-hot encoding for categoricals, and scaling for LR/SVC/MLP) was fit within each training fold to avoid leakage. Performance is reported as mean AUROC. The LightGBM configuration with the highest outer-fold AUROC was selected as the baseline.For the ensemble model, We first trained a LightGBM classifier on the morphometric features extracted from the pathology reports. In parallel, we obtained the 768-dimensional `[CLS]` embeddings from the trained CLIP alignment model, truncated them to the first 256 dimensions, and then applied ℓ_2_ normalization (unit length) to each embedding vector. A Support Vector Classifier(SVC) was fitted on these normalized 256-dimensional embeddings to predict the complication outcome. The final per-patient probability was computed by soft voting with fixed equal weights (1:1) between the morphometric LightGBM and the [CLS]–based SVC model.

### 4.5 Subgroup Analysis

Subgroups were defined a based on clinical relevance: (1) age *<* 65 vs. ≥ 65 years; (2) tissue expander–to–implant interval (EII; coded as `TEtoImplant`) *<* 180 vs. ≥ 180 days; and (3) radiation timing with four levels: none, before reconstruction, during staged reconstruction, and after reconstruction.

All subgroup analyses used out-of-fold (OOF) patient-level predictions from the outer cross-validation. The primary endpoint was discrimination (AUROC). As a secondary endpoint, we computed sensitivity at 80% specificity (sens@80). For sens@80, within each evaluation we selected the threshold that achieved a false-positive rate closest to 0.20.

We formed confidence intervals (CI) from the pooled OOF predictions and reference labels for each subgroup. For a given subgroup, we repeatedly (B = 2000) drew stratified bootstrap samples of the OOF rows—sampling positives and negatives separately with replacement so that each replicate kept the original class counts. Model scores were fixed (no refitting); in every replicate we recomputed the subgroup AUROC on the resampled pairs. The 2.5th and 97.5th percentiles of the bootstrap AUROC distribution yielded the two-sided 95% CI. Note that the resampling operates on the concatenated OOF set, so fold membership is not re-imposed during the bootstrap.

### 4.6 Feature-level interpretation & interaction

SHAP analyses were performed on the *tree-based* component of the ensemble (Light-GBM over raw clinical and weight features). We restricted attributions to this branch because TreeExplainer provides faithful, low-variance explanations for gradient-boosted trees, whereas the parallel SVC branch operates on high-dimensional CLS embeddings that lack per-dimension clinical semantics.

We computed feature attributions with the Python `shap` package (TreeExplainer) on the LightGBM submodel of the final model (baseline + both CLS). Explanations were run on the full dataset passed through the model’s own preprocessing. We visualized the top–10 features via a mean-|SHAP| bar plot and a matching beeswarm; dependence/interaction figures used `shap.dependence plot`.

## 5 Data Availability

The de-identified structured clinical dataset is available from the corresponding author upon reasonable request. To protect patient privacy, the free-text pathology reports are not publicly available but may be shared with qualified researchers following the execution of a Data Use Agreement (DUA) and institutional approval.

## 6 Ethics statement

The Institutional Review Board of Cedars-Sinai Medical Center gave ethical approval for this work.

**Table S1:** Baseline demographic and clinical characteristics of the study cohort (*N* = 963).

| Characteristic | Value (%) |
| --- | --- |
| Age, years (mean ± SD) | 56.3 ± 11.6 |
| BMI, kg/m<sup>2</sup> (mean ± SD) | 24.9 ± 4.8 |
| Underweight | 33 (3.5) |
| Healthy weight | 524 (55.5) |
| Overweight | 259 (27.4) |
| Obesity | 128 (13.6) |
| Laterality |  |
| Unilateral | 194 (20.2) |
| Bilateral | 769 (79.8) |
| Smoking status |  |
| Current smoker | 38 (3.9) |
| Former smoker | 160 (16.7) |
| Never smoker | 765 (79.4) |
| Diabetes status |  |
| Has Diabetes | 64 (6.7) |
| No Diabetes | 899 (93.3) |
| Radiation timing |  |
| None | 717 (74.4) |
| Pre-reconstruction | 72 (7.5) |
| Mid-reconstruction | 127 (13.2) |
| Post-reconstruction | 47 (4.9) |
| Chemotherapy |  |
| Neoadjuvant | 103 (10.7) |
| Adjuvant | 127 (13.2) |
| None | 733 (76.1) |
| Implant location |  |
| Subpectoral | 739 (76.7) |
| Prepectoral | 224 (23.3) |

**Table S2:** Different model performance and pretrained embeddings performance(*n* = 10 folds).

| Model | AUROC (SD) |
| --- | --- |
| LightGBM | 0.740 (50) |
| LogReg (l1) | 0.687 (57) |
| LogReg (l2) | 0.691 (54) |
| RF | 0.736(70) |
| SVC | 0.722 (60) |
| MLP | 0.673 (52) |
| Gemma Embeddings (control) | 0.750 (43) |

**Table S3:** Cross-modal retrieval on the validation set (*N* =215). Chance baseline is *K/N*.

| Direction | R@1 | R@5 | R@10 | R@20 | R@50 |
| --- | --- | --- | --- | --- | --- |
| Clinical→Pathology | 0.074 | 0.288 | 0.409 | 0.581 | 0.781 |
| × chance (K/N) | 16.0× | 12.4× | 8.8× | 6.25× | 3.36× |
| Pathology→Clinical | 0.088 | 0.279 | 0.414 | 0.554 | 0.772 |
| × chance (K/N) | 19.0× | 12.0× | 8.9× | 5.95× | 3.32× |

**Table S4:** Subgroup performance by comorbidities and smoking status (condition present). Metrics are computed from out-of-fold predictions of the best model. sens@80 = sensitivity at 80% specificity; spec@80 = specificity at 80% sensitivity.

| Subgroup (condition present) | $N_{all}$ | $N_{pos}$ | $N_{neg}$ | AUC | sens@80 | spec@80 | acc |
| --- | --- | --- | --- | --- | --- | --- | --- |
| Current smoker | 31 | 10 | 21 | 0.847 | 0.800 | 0.905 | 0.871 |
| Hypertension | 163 | 71 | 92 | 0.781 | 0.606 | 0.587 | 0.755 |
| Thyroid disorder | 118 | 41 | 77 | 0.772 | 0.634 | 0.597 | 0.771 |
| Hyperlipidemia | 129 | 45 | 84 | 0.780 | 0.667 | 0.500 | 0.783 |

